# Are socio-demographic factors associated with electronic Health Information System utilization? *A cross sectional study of the District Health Information System -2 (DHIS-2) in Burundi*

**DOI:** 10.1101/2024.01.03.24300804

**Authors:** Innocent Yandemye, Alexandre Nimubona

**Affiliations:** Health policy advisor, Ministry of Health, Bujumbura, Burundi; Independent Researcher, Burundi country, Burundi

## Abstract

The utilization of Electronic Health Information System is crucial in informing credible decision-making process, especially in developing countries. We aimed to investigate the socio-demographic factors of Health workers that are associated with their utilization of the DHIS-2 platform in Burundi. We conducted a cross-sectional study amongst health workers involved in collecting and validating data in health facilities. The male gender was more associated with the DHIS-2 effect on the enhancement of health information access (χ2 = 7.995, P=.005) than the female gender. Marital status was associated with quality information (χ2 = 6.437, P= .011) and health information management (χ2= 5.053, P= .025) improvement. The lower time travel to workplace was associated with the improvement of quality information (χ2= 11.224, P= .001) and health information management (χ2= 6.568, P= .010), training on DHIS-2 (χ2= 16.374, P= .000), DHIS-2 compatibility with health facility management tools (χ2 = 4.726, P= .030), national health information system design (χ2= 8.023, P= .005), compatibility with primary health information tools (χ2= 9.339, P= .002) and the national health policy objective (χ2= 9.699, P= .002), required security elements (χ2= 14.205, P= .000), data transfer security (χ2= 6.288, P= .012), data protection against modification (χ2= 6.497, P= .011), and assurance of personal privacy (χ2= 7.650, P= .006). The lower educational level was associated with information error minimization (χ2= 7.243, P= .007) and learning ease (χ2 = 4.175, P= .041). The health worker function was associated with DHIS-2 compatibility with national health policy objective (χ2 =4.496, P= .034). The geographical area of work was associated with DHIS-2 ease of use and interoperability assurance (χ2 = 10.287, P= .006; χ2 = 16.086, P= .000). There is a need to make the use of the DHIS-2 platform a global public health concern in fragile health systems for robust decision-making process.

## Introduction

There is a global consensus that the Health Information System (HIS) plays a pivotal role in advancing Universal Health Coverage (UHC) [1, 2]. According to Sunny *et al*.[3], the HIS requires resources and regular monitoring of health data from the points of care to national level in order to inform decision making process for sustainable health outcomes in countries [3].

In this regard, the HIS is transitioning from paper-based to electronic-based, with a view to comply with the rapid evolution of health technology for UHC, especially in developing countries[4]. In global public health perspective, the electronic-based HIS(eHIS) has been proven as a means to improve quality data while aligning fragmented reporting demands from bilateral and multilateral funding agencies, and policy makers[5, 6]. Evidence suggests that the utilization of eHIS is crucial in informing credible decision-making for UHC[7].

Currently, many developing countries are implementing DHIS-2 as electronic-based data management system to achieve UHC[8]. Worryingly, strong emphasis has been placed at national, regional, and provincial levels – Many technical health workers in both health district offices and health facilities who are supposed to significantly contribute in the use and operationalization of the DHIS-2 platform are left behind[9, 10].

In Burundi, the DHIS-2 platform has been introduced in the health system since 2014 in order to strengthen the health system in data management and respond to both National Health Policy and National Plan for Health Development [11]. Regardless of the financial and technical efforts of the Ministry of Health and funding agencies to operationalize the DHIS-2 platform at local levels, the HIS is still facing many challenges such as: (i) utilization of paper-based system when collecting data in the health district offices and health facilities – this leads to several number of mistakes in data reporting system; and (ii) the lack of connection or network for data entry and reporting in such offices and facilities. This is compromising health data quality for robust UHC-related decision making process [12, 13].

Extensive research has shown that involving technical health workers in using and operationalizing the DHIS-2 platform for health data management can lead to expected health outcomes [14–18]. It remains unclear whether socio-demographic factors may influence such outcomes – to our knowledge, no studies are interested in learning the association between socio-demographic factors and the use of the DHIS-2 platform at this moment.

The aim of this study was to investigate the socio-demographic factors of health workers that are associated with their utilization of the DHIS-2 platform in Burundi.

## Materials and methods

### Study setting

This study was conducted in Burundi, a country with fragile health system. The DHIS-2 is an essential resource that Burundi is adopting to transition the health system to digitalization and accelerate the achievement of UHC. To do so, Burundi intends to train key health system staff on DHIS-2 utilization and provide them with the necessary technology to collect and analyze health data in order to promote evidence-informed decision making process for UHC, from community to national levels[4]. We are interested in confirming or infirming this null hypothesis: “there is no relationship between socio-demographic factors and the use of the DHIS-2 platform in Burundi.”

### Study design

We conducted a cross-sectional study amongst health workers involved in collecting and validating data in health facilities. We chose this design for two major reasons. First, the design is usually used for population-based surveys and is quite inexpensive. Second, a cross-sectional survey proved to be easy to conduct[19]. We have no particular reason for framing our survey duration – we stopped the survey after we had reached the entire expected population of our study. This took four months, from July 3 to October 29, 2023. However, one year before a decade of DHIS-2 platform utilization in Burundi [4], we thought that it was vital to learn about socio-demographic factors that can influence the utilization of such a platform.

### Study population and sampling methods

The study population was the health workers that are involved in health data collection and validation at the points of care. We used a multi-stage sampling method to identify and select participants from the study population as indicated below.

#### Stage 1

We purposively chose three of Burundi’s 18 health provinces based on their geographical location to ensure that health workers from rural and urban provinces of the country were represented. Kirundo and Cankunzo were chosen because, according to our knowledge of the context, they are the most geographically remote provinces in Burundi, with limited internet access to operationalize the DHIS-2 platform. The two provinces may represent health workers from remotest areas. Muramvya province was chosen because it is located between the political and economic capital of the country – the province may be representative of health workers from both urban and semi-urban areas in Burundi.

#### Stage 2

Census-based sampling of health district offices – the three provinces have respectively two, four, and two health district offices, which were all included in the study. In total the study included eight health district offices.

#### Stage 3

The eight health districts have a total of 137 health facilities (public and private), of which we selected 76 at random. We excluded 44 private health facilities from the sample because they frequently have no publicly accessible data[20] – this should decrease the robustness of our findings. We used the sample calculator (http://www.raosoft.com/samplesize.html) to determine the sample size of 76 from the population size of 93, applying a confidence level of 95%. We established a list of the 93 health facilities and then used the Random Number Table to systematically pick the 76 health facilities.

#### Stage 4

In each of the 76 health facilities, we chose one participant who was available during the data collecting period between the data and health facility manager. Furthermore, we recruited a DHIS-2 manager from each of the eight health district offices because they are directly involved in data validation in health facilities. At the end, 84 health workers took part in our study.

### Data collection

We applied the survey method to collect data because it was most appropriate for our study design [19]. After a pilot survey of 20 health data managers picked by convenience from 20 health facilities outside of our study sites, we used the Cronbach’s alpha test (0.809) to validate our questionnaire. The latter was also validated by DHIS-2 experts working in the department of the National Health Information System in Burundi. Data were collected at a single point in time, with no follow-up. Participants were found at capacity-building meetings organized by the Ministry of Health at the health district level. We applied the technique of self-administered survey to distribute the questionnaire in-person. Respondents filled the questionnaire in the presence of the researcher to answer any clarification questions and check that all items were responded to. The questionnaire was in French, which is the official language used in Burundi. We translated the French version of transcribed data in English.

### Measurement variables

We measured four groups of dependent variables termed DHIS-2 management tools[21, 22].

First, there is a group of six variables related to **administrative support** including internet availability; financial resource allocation in DHIS-2 by health facility; health facility engagement with DHIS-2 functionality; health facility encouragement on DHIS-2 utilization, availability of technical support; and satisfaction with DHIS-2 training.

Second, the group of eleven variables related to **DHIS-2 design** comprising required security elements; data transfer security; data protection against modification; assurance of personal privacy; learning ease; interoperability assurance; ease of use; compatibility with primary health information tools such as patient records and registers; compatibility with current health facility management tools; national health information system design; and compatibility with national health policy objective.

Third, a group of five variables connected to **DHIS-2 effect on data utilization** such as efficiency and effectiveness of operations; improvement of quality information, information management improvement; enhancement of health information accessibility; and information error minimization.

Fourth, a group of four variables linked with **services provision** that consist of quality services improvement; increase of health services volume; operational cost reduction; and efficiency enhancement.

Independent variables included two groups of socio-demographic characteristics(factors) of health workers. There is a group of five demographic characteristics comprising gender, marital status, age, educational level, and geographical area of work (remote and urban or semi-urban areas expressed here in terms of province). The other is a group of professional characteristics including years with DHIS-2 utilization, place of work (facility level and health district office), time travel to service, and function of the health worker (health facility or data manager). These variables were chosen based on the study of Jamebozorgi *et al*. [23].

### Data analysis

The latest version of SPSS (version 25), was used to organize and analyze data. We utilized Chi-Square test to measure the association between the four groups of dependent variables (related to the DHIS-2 platform utilization) and the socio-demographic factors described above. Based on the alpha level of 0.05, we used the results from Chi-Square test to estimate the degree of significance and decide whether to reject or keep the null hypothesis.

### Ethics

After explaining the goal of the study, every participant was asked to give oral and written consent to participate. For those who accepted to participate, the following statement was written at the beginning of the questionnaire sheet: “I accept to take part in this study. I acknowledge that I am aware of the purpose of this study and that I am over 18 years of age.”

Every respondent jointly signed the fulfilled questionnaire with the researcher, which also included informed consent. The researcher took the original filled questionnaire, whereas the respondent kept a copy of the original filled questionnaire.

The General Directorate of Health Services Provision and Accreditations granted the study the ethical approval (N/Réf: 635/52/DGOSA/2023).

## Results

Table 1 indicates categories and frequencies of sociodemographic factors studied.

**Table 1.** Socio-demographic factors.

| Variables | Categories | Numbers | Frequency (%) |
| --- | --- | --- | --- |
| Gender | Male | 69 | 82.1 |
|  | Female | 15 | 17.9 |
| Marital status | Married | 69 | 82.1 |
|  | Single | 15 | 17.9 |
| Age in years | <=40 | 71 | 84.5 |
|  | >40 | 13 | 15.4 |
| Years with DHIS-2 utilization | <=5 | 63 | 75 |
|  | >5 | 21 | 25 |
| Place of work | Health center | 67 | 79.8 |
|  | Hospital | 9 | 10.7 |
|  | District office | 8 | 9.5 |
| Time travel to service in hour | ≤1 | 57 | 67.8 |
|  | >1 | 27 | 32.1 |
| Province | Cankuzo | 25 | 29.7 |
|  | Kirundo | 30 | 35.7 |
|  | Muramvya | 29 | 34.5 |
| Educational level | High school diploma | 56 | 66.7 |
|  | University level | 28 | 33.3 |
| Health worker function | Health facility manager | 19 | 22.6 |
|  | Data manager | 65 | 77.4 |

Eighty-four participants from 76 health facilities and 8 health district offices responded to the questionnaire. Most of them were male (82.1%), married (82.1%), less than or equal to 40 years of age (84.5%), less than or equal to 5 years of experience (75%), data manager (77.4%), from health centers (79.8%), with a travel time to service of equal or less than one hour (67.8%), from Kirundo province (35.7%), and with a secondary level of education (66.7%).

### Chi-square test results for independent and dependent variables

We decided to report only statistically significant results in order to confidently visualize the influence of our factors of interest, which resulted in the rejection of the aforementioned null hypothesis.

**Table 2.**
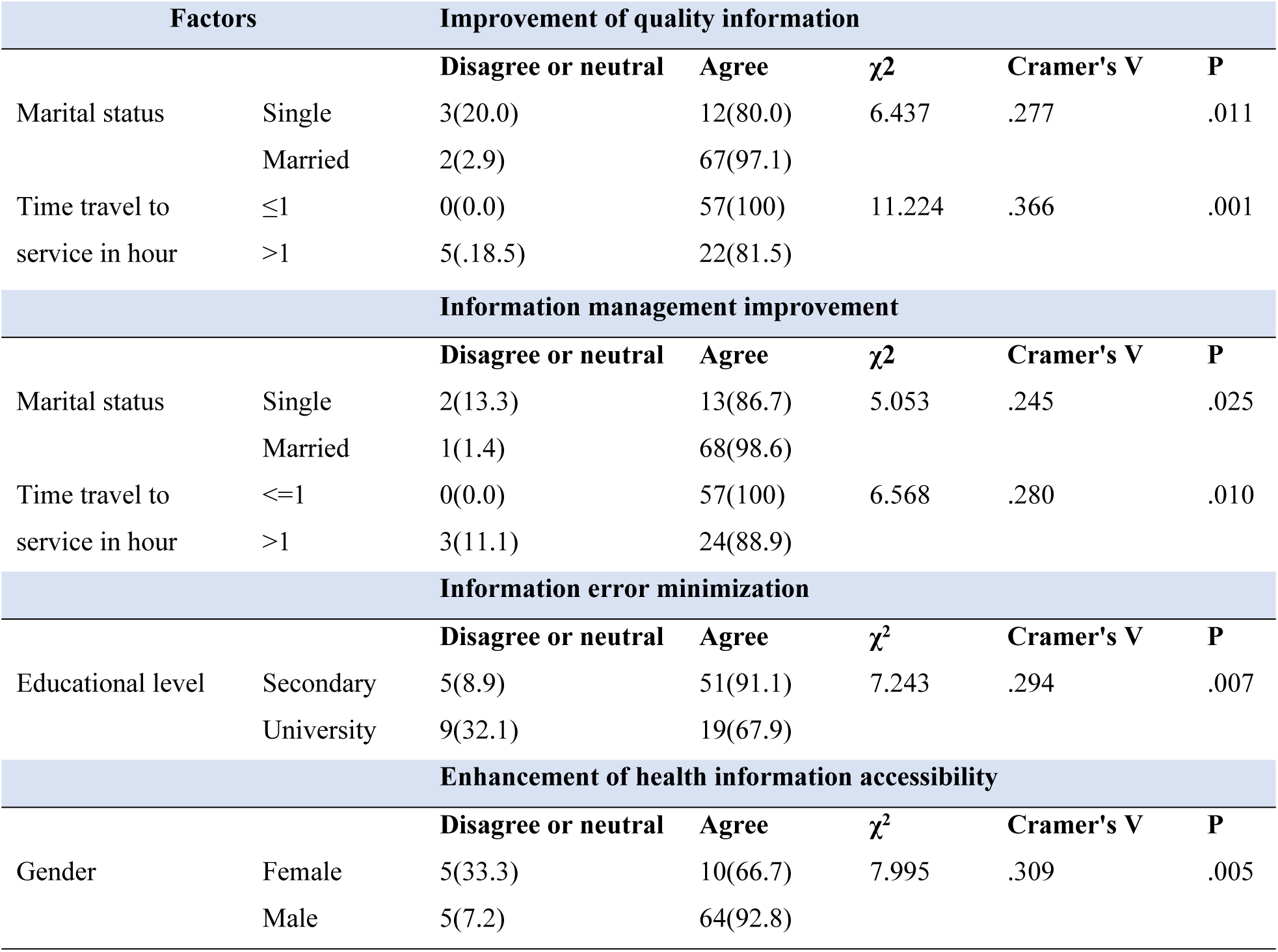
Chi-square test of socio-demographic factors and DHIS-2 effect on data utilization.

Marital status and time travel to service were significantly associated with both improvement of quality information (χ^2^= 6.437, P= .011 and χ^2^= 11.224, P= .001) and information management (χ^2^= 5.053, P=.025 and χ^2^ = 6.568, P=.010). For this improvement of quality information and health information management, the marital status included 97.1% and 80.0% of married participants, and 98.6% and 86.7% of single participants, respectively.

The time travel to service included 100% of participants taking equal or less than one hour to reach the point of service, whereas participants taking more than one hour to reach the point of service represented 81.5% for the improvement of quality information and 88.9% for the improvement of the health information management.

Gender was significantly associated with the enhancement of health information accessibility (χ^2^= 7.995, P=.005), with 92.8% for males and 66.7% for females.

Information error minimization was significantly associated with educational level (χ^2^ = 7.243, P=.007), with 91.1% for a secondary level and 67.9% for university level.

**Table 3.** Chi-square test analysis of socio-demographic factors and DHIS-2 administrative support.

|  |  | Satisfaction with DHIS-2 training |  |  |  |  |
| --- | --- | --- | --- | --- | --- | --- |
| Factor | | Disagree or Neutral | Agree | $\chi^2$ | Cramer's V | P |
| Time travel to service in hour | $\leq 1$ | 2(42.1) | 33(57.9) | 16.374 | .442 | .000 |
| | $> 1$ | 24(88.9) | 3(11.1) | | | |

The time travel to service was significantly associated with satisfaction on DHIS-2 training (χ2 = 16.374, P= .000). Participants with a time travel to service of one hour or less represented 57.9% whereas participants with a time travel to service of more than one hour represented 11.1%.

**Table 4.**
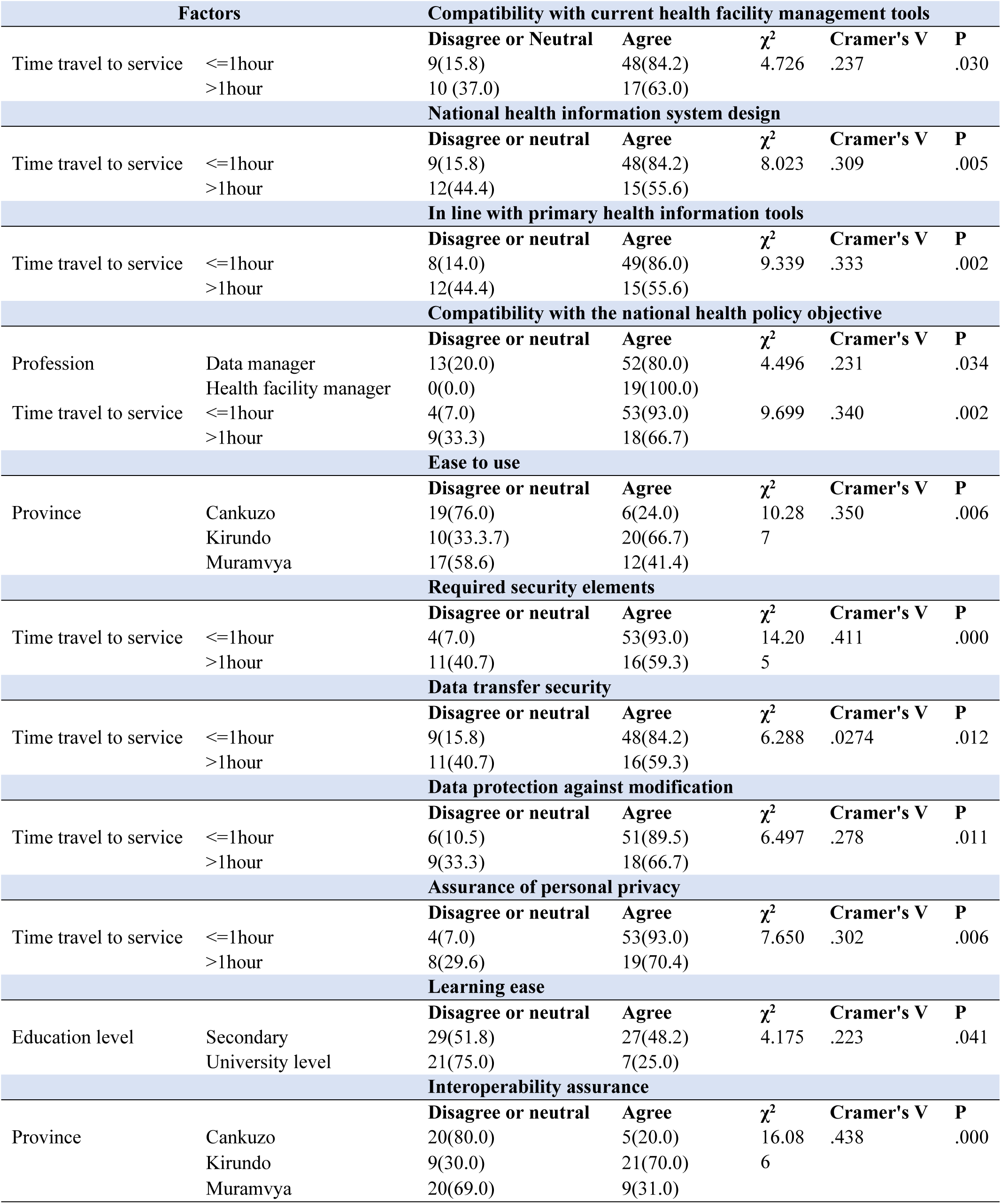
Chi-square test analysis of socio-demographic factors and DHIS-2 design.

The time travel to service was significantly associated with: (i)DHIS-2 compatibility with health facility management tools (χ2 = 4.726, P=.030), (ii)national health information system design (χ2 = 8.023, P=.005), (iii)compatibility with primary health information tools such as patient records and registers (χ2 = 9.339.023, P=.002), (iv)compatibility with the national health policy objective (χ2= 9.699, P=.002), (v)required security elements (χ2= 14.205, P=.000), (vi)data transfer security (χ2= 6.288, P=.012), (vii)data protection against modification (χ2= 6.497, P=.011),and (viii)assurance of personal privacy (χ2 = 7.650, P=.006). Participants with a time travel equal to or less than one hour represented respectively 84.2%, 84.2%, 86.0%, 93.0%, 93.0%, 84.2%, 89.5%, and 93.0%.

Health worker function was significantly associated with the DHIS-2 compatibility with the national health policy objective (χ2=4.496, P=.034) in 100% of health managers and 80% of data managers and the geographical area of work (province) was associated with DHIS-2 ease of use and interoperability assurance (χ2= 10.287, P= .006 and; χ2 = 16.086, P= .000). The remotest area represented respectively 66.7% and 70% (Kirundo province), 24.0% and 20.0% (Cankuzo province). The urban and semi-urban areas represented respectively 41.4 % and 31.0% (Muramvya province). Educational level was significantly associated with learning ease (χ2 = 4.175, P=.041), the secondary level represented 48.2%.

## Discussion

To our knowledge, this is the first study exploring the socio-demographic factors associated with the use of the DHIS-2 platform in Burundi. It taken place in three of the country’s 18 provinces. The Majority of the 84 participants were male (82.1%) amongst them, 79.8% came from health centers. Similarly, a study in Sierra Leone found that 77% of DHIS-2 users were male[21]. In contrast, a Brazilian study found that females were the most frequent users, with no relation to socio-economic factors [24].

The male gender was more associated with the DHIS-2 effect on the enhancement of health information access (χ2 = 7.995, P=.005) than the female gender. These findings are consistent with that of a previous study conducted in Ethiopia, where male gender was significantly associated with knowledge of electronic health records [25]. In contrast, a study in Nigeria found the female gender significantly associated with the use of electronic health information [26] – surprisingly, the DHIS-2 necessitates regular use of computers and the internet, and a large body of literature indicates that the males use computers and the internet more frequently than females[27, 28].

Marital status was significantly associated with both improvement of quality information (χ2 = 6.437, P= .011) and health information management(χ2= 5.053, P= .025). These findings tie with previous studies in the USA, wherein being married was significantly associated with the use of digital health interventions[29]. Two major reasons could explain this observation. First, it seems possible that married health workers have a more stable place of residence than single ones – this may give married health workers a sense of stability and emotional support, which may have a positive influence on their services. Second, the increased responsibilities of married health workers in the family, may increase the perceived value of their job.

The lower time travel to reach health workplace was significantly associated with the improvement of quality information (χ2= 11.224, P= .001) and health information management (χ2= 6.568, P= .010), training on DHIS-2 (χ2= 16.374, P= .000), DHIS-2 compatibility with health facility management tools (χ2 = 4.726, P= .030), national health information system design (χ2= 8.023, P= .005), compatibility with primary health information tools such as records and patient registers (χ2= 9.339, P= .002) and the national health policy objective (χ2= 9.699, P= .002), required security elements (χ2= 14.205, P= .000), data transfer security (χ2= 6.288, P= .012), data protection against modification (χ2= 6.497, P= .011), and assurance of personal privacy (χ2= 7.650, P= .006). These results reflect those reported in a study conducted in West Asia, which found that short time travel from home to the point of primary healthcare services has increased health workers performance and satisfaction [30]. According to a study conducted in America amongst 1044 physicians, the use of electronic health records has significantly increased work at home overtime – so, it is possible that the use of DHIS-2 can also extend work at home: health workers living nearest the point of service reach their home early and gain more time to explore DHIS-2[31].

Our study found that lower educational level was higher significantly associated with information error minimization (χ2= 7.243, P= .007) and learning ease (χ2 = 4.175, P= .041) compared to higher educational level. These findings corroborate the results of previous studies conducted in Turkey and Albania, which indicated that low education level was associated with job satisfaction[32,33].Research conducted in developed countries have found that a higher education level is associated with higher e-health utilization [34–36]. Another study conducted in Burundi on satisfaction with performance-based financing(PBF) has found that lower education level was associated with error minimization in electronic-based data reporting[37]. Our findings can be explained by the low wage levels observed amongst health workers with a high level of education in Burundi. In the same line, experience in developing countries has demonstrated that higher education increases the demand for higher-level positions and considerations amongst health workers [35]. When that demand is not satisfied, it can lead to work demotivation [38, 39]. According to our experiences in the Burundi health system, there is no clear separation of task attribution amongst health workers with different levels of education. For example, health workers with secondary education have the same roles and responsibilities as health workers with university degrees in eHIS management. Furthermore, there is no clear distinction between general and specialist doctors in terms of health data generation (i.e., specialized consultations are most of the time devoted to general doctors). This could explain the increased level of significance of information error minimization reported in health workers with lower education levels compared to those with higher education levels.

The health worker function was significantly associated with DHIS-2 compatibility with the national health policy objective (χ2 =4.496, P= .034). These findings are in line with previous findings from Asia that show a significant association between health workers occupation and electronic health utilization[40,41]. Our results can be explained by the differences in understanding the national health policy and the vision of the Ministry of Health amongst health workers. Experience has shown that health facility managers are more likely to participate in multiple trainings organized by the Ministry of Health and funded by its funding agencies, gaining knowledge of the Ministry of Health’s national health policy and vision, whereas other health workers receive limited trainings focused to their daily tasks.

Another interesting finding was that the geographical area of work (province) was significantly associated with DHIS-2 ease of use and interoperability assurance (χ2 = 10.287, P= .006; χ2 = 16.086, P= .000) with high rates in Kirundo province – 66.7% and 70%, respectively. Kirundo Province was one of the provinces that piloted DHIS-2 in the Burundi health system. The higher significance reported in this province can thus be explained by its extended experience with DHIS-2 utilization in comparison to the other provinces studied.

### Limitations

This study mainly used quantitative methods for data collection and analysis. We are convinced that using only quantitative information cannot lead to solid inferences tailored to the context of the Burundi health system. Qualitative studies focusing on the behaviors of both health workers and health system management should provide additional information that can go beyond the scope of the current study.

## Conclusion

Our study indicated that a significant number of socio-demographic factors are associated with the use of DHIS-2 platform in the Burundi health system. This association is more significant in remotest areas with long experience of DHIS-2 operationalization compared to urban and semi-urban areas. Also, the higher level of education was negatively associated with the use of DHIS-2 platform. Given that decisions are made by highly educated health workers (only medical doctors in the Burundi health system), this may jeopardize the quality of the decision-making process for universal health coverage. The other important learning from our study is that the use of DHIS-2 platform increases time spent at home by health workers in charge of eHIS management. This implies that the use of DHIS-2 can promote telework, which is one of the strategies that the Burundi health system can use to incentivize eHIS managers and enhance their wellbeing by working from home for a set period of time. In sum, there is a need to make the use of the DHIS-2 platform a global public health concern in fragile health systems for robust decision making process to achieve Universal Health Coverage.

## Data Availability

Data are accessible upon reasonable request. All relevant data are included in the paper. Readers interested can contact the corresponding author for access to the data.

## Acknowledgements

To the Ministry of Health of Burundi, especially the General Directorate of Health Services and the Directorates of Kirundo, Cankuzo and Muramvya Health Provinces, for granting permission to collect data. To the health facility and district managers for facilitating the data collection process.

## Author contributions

**Conceptualization**: Innocent Yandemye, Alexandre Nimubona

**Data curation** : Innocent Yandemye

**Formal analysis** : Innocent Yandemye

**Funding acquisition**: Not applicable

**Investigation**: Innocent Yandemye

**Methodology**: Alexandre Nimubona, Innocent Yandemye

**Project administration**: Innocent Yandemye

**Resources**: Innocent Yandemye, Alexandre Nimubona

**Software**: Innocent Yandemye

**Supervision**: Innocent Yandemye

**Validation** : Alexandre Nimubona

**Visualization** : Innocent Yandemye, Alexandre Nimubona

**Writing – original draft** : Innocent Yandemye

**Writing – review & editing:** Alexandre Nimubona

